# Increased mortality among individuals hospitalised with COVID-19 during the second wave in South Africa

**DOI:** 10.1101/2021.03.09.21253184

**Authors:** Waasila Jassat, Caroline Mudara, Lovelyn Ozougwu, Stefano Tempia, Lucille Blumberg, Mary-Ann Davies, Yogan Pillay, Terrence Carter, Rams Morewane, Milani Wolmarans, Anne von Gottberg, Jinal N. Bhiman, Sibongile Walaza, DATCOV Author Group, Cheryl Cohen

**Affiliations:** National Institute for Communicable Diseases (NICD) of the National Health Laboratory Service (NHLS), Johannesburg, South Africa; School of Public Health, Faculty of Health Sciences, University of Witwatersrand, Johannesburg, South Africa; Western Cape Government: Health, Health Impact Assessment Directorate, Cape Town, South Africa; Clinton Health Access Initiative, Pretoria, South Africa; National Department of Health, Pretoria, South Africa; School of Pathology, Faculty of Health Sciences, University of the Witwatersrand

**Keywords:** COVID-19, hospital admissions, mortality, first and second wave, new variant

## Abstract

**Introduction:** South Africa experienced its first wave of COVID-19 peaking in mid-July 2020 and a larger second wave peaking in January 2021, in which the SARS-CoV-2 501Y.V2 lineage predominated. We aimed to compare in-hospital mortality and other patient characteristics between the first and second waves of COVID-19.

**Methods:** We analysed data from the DATCOV national active surveillance system for COVID-19 hospitalisations. We defined four wave periods using incidence risk for hospitalisation, pre-wave 1, wave 1, pre-wave 2 and wave 2. We compared the characteristics of hospitalised COVID-19 cases in wave 1 and wave 2, and risk factors for in-hospital mortality accounting for wave period using multivariable logistic regression.

**Results:** Peak rates of COVID-19 cases, admissions and in-hospital deaths in the second wave exceeded the rates in the first wave (138.1 versus 240.1; 16.7 versus 28.9; and 3.3 versus 7.1 respectively per 100,000 persons). The weekly average incidence risk increase in hospitalisation was 22% in wave 1 and 28% in wave 2 [ratio of growth rate in wave two compared to wave one: 1.04, 95% CI 1.04-1.05]. On multivariable analysis, after adjusting for weekly COVID-19 hospital admissions, there was a 20% increased risk of in-hospital mortality in the second wave (adjusted OR 1.2, 95% CI 1.2-1.3). In-hospital case fatality-risk (CFR) increased in weeks of peak hospital occupancy, from 17.9% in weeks of low occupancy (<3,500 admissions) to 29.6% in weeks of very high occupancy (>12,500 admissions) (adjusted OR 1.5, 95% CI 1.4-1.5).

Compared to the first wave, individuals hospitalised in the second wave, were more likely to be older, 40-64 years [OR 1.1, 95% CI 1.0-1.1] and ≥65 years [OR 1.1, 95% CI 1.1-1.1] compared to <40 years; and admitted in the public sector [OR 2.2, 95% CI 1.7-2.8]; and less likely to have comorbidities [OR 0.5, 95% CI 0.5-0.5].

**Conclusions:** In South Africa, the second wave was associated with higher incidence and more rapid increase in hospitalisations, and increased in-hospital mortality. While some of this is explained by increasing pressure on the health system, a residual increase in mortality of hospitalised patients beyond this, could be related to the new lineage 501Y.V2.

**RESEARCH IN CONTEXT:** Evidence before this study
Most countries have reported higher numbers of COVID-19 cases in the second wave but lower case-fatality risk (CFR), in part due to new therapeutic interventions, increased testing and better prepared health systems. South Africa experienced its second wave which peaked in January 2021, in which the variant of concern, SARS-CoV-2 501Y.V2 predominated. New variants have been shown to be more transmissible and in the United Kingdom, to be associated with increased hospitalisation and mortality rates in people infected with variant B.1.1.7 compared to infection with non-B.1.1.7 viruses. There are currently limited data on the severity of lineage 501Y.V2.

Added value of this study
We analysed data from the DATCOV national active surveillance system for COVID-19 hospitalisations, comparing in-hospital mortality and other patient characteristics between the first and second waves of COVID-19. The study revealed that after adjusting for weekly COVID-19 hospital admissions, there was a 20% increased risk of in-hospital mortality in the second wave. Our study also describes the demographic shift from the first to the second wave of COVID-19 in South Africa, and quantifies the impact of overwhelmed hospital capacity on in-hospital mortality.

Implications of all the available evidence
Our data suggest that the new lineage (501Y.V2) in South Africa may be associated with increased in-hospital mortality during the second wave. Our data should be interpreted with caution however as our analysis is based on a comparison of mortality in the first and second wave as a proxy for dominant lineage and we did not have individual-level data on lineage. Individual level studies comparing outcomes of people with and without the new lineage based on sequencing data are needed. To prevent high mortality in a potential third wave, we require a combination of strategies to slow the transmission of SARS-CoV-2, to spread out the peak of the epidemic, which would prevent hospital capacity from being breached.

## INTRODUCTION

South Africa reported its first case of PCR-confirmed SARS-CoV-2 infection on 5 March 2020, and since then has experienced a first wave peaking in mid-July and a larger second wave peaking in early January 2021.(1) A new lineage of SARS-CoV-2 detected in the Eastern Cape province in September (2) was then reported to be the predominant lineage of initially-tested samples in each of the Eastern Cape (154/157, 98%), Western Cape (137/174, 79%), KwaZulu-Natal (244/252, 97%) and Gauteng (126/150, 84%) provinces between December 2020 and February 2021.(3,4) This lineage, named 501Y.V2, contains several mutations that were not identified in SARS-Cov-2 viruses from South Africa prior to September 2020. Eight of these mutations are in the spike protein coding region and have been shown to enhance binding to the human ACE2 receptor (5).

Preliminary data suggest that SARS-CoV-2 501Y.V2 may be more transmissible than other lineages (2,6) and escape the immune response from preceding infection with old SARS-CoV-2 lineages. (7,8) The United Kingdom reported increased hospitalisation and mortality rates in people infected with variant B.1.1.7 compared to infection with non-B.1.1.7 viruses. (9,10) There are currently limited data on the severity of lineage 501Y.V2. It is important to determine the severity of disease during our second wave with 501Y.V2 lineage predominance, to better understand the burden of COVID-19 mortality and impact on health care services.

We aimed to describe, among hospitalised individuals with laboratory-confirmed COVID-19 throughout South Africa, the demographic and clinical characteristics of individuals in the first and second waves and assess risk factors for in-hospital mortality.

## METHODS

### Setting

South Africa, a temperate country in the Southern Hemisphere, has recorded the most COVID-19 cases in Africa. (11) The country has a high burden of non-communicable diseases and obesity, HIV and tuberculosis. According to the 2016 Demographic and Health Survey 41% of adult women and 11% of men were obese, 46% of women and 44% of men had hypertension, and 13% of women and 8% of men had diabetes. (12) In 2019, there were 7.5 million people estimated to be living with HIV in South Africa, of which 2.3 million (31%) were not receiving treatment. (13) In 2018, 301,000 new cases of TB were diagnosed in South Africa. (14) South Africa has a dual health system with a publicly funded district health system, that serves approximately 84% of the population, and a private health system largely funded by private health insurance schemes. (15)

### Data collection

Secondary data analysis was conducted of the DATCOV database between 5 March 2020 and 6 February 2021. DATCOV is an active surveillance system for COVID-19 admissions established in March 2020, that has achieved comprehensive coverage of all hospitals in South Africa which have admitted a COVID-19 case. (16) As of 6 February 2021, a total of 627 facilities submitted data on hospitalised COVID-19 cases, 377 from the public sector and 250 from the private sector. DATCOV contains data on all individuals, who had a positive real-time reverse transcription polymerase chain reaction (rRT-PCR) assay for SARS-CoV-2 or a person who had a positive SARS-CoV-2 antigen test, with a confirmed duration of stay in hospital of one full day or longer, regardless of age or reason for admission. The case reporting form was adapted from the World Health Organization (WHO) COVID-19 case reporting tool and contains the following variables: basic demographic data, exposures such as occupation, and potential risk factors such as obesity, comorbid disease(s), and pregnancy status. Additional variables included data on level of treatment (ward, high dependency, or intensive care unit), complications, treatment, and outcomes of the hospital admission (discharged, transferred out to another hospital, or died). Data collection was either through direct entry onto the DATCOV online platform, or through import of electronic data from health information systems into the database. Data imports contained validation checks to identify data errors.

Case-fatality risk (CFR) was calculated among individuals with in-hospital outcome, *i*.*e*. COVID-19 deaths divided by COVID-19 deaths plus COVID-19 discharges, excluding individuals who are still admitted in hospital. For the calculation of incidence risks, Statistics South Africa mid-year population estimates for 2020 were utilised. (17)

### Selection of time period for analysis

The periods of analysis were estimated using hospital admission data in the country. The wave period was defined from the time the country recorded a weekly incidence risk of 5 admissions per 100,000 population at the start of the wave to the same incidence risk at the end of the wave. For the analysis of factors associated with in-hospital mortality the COVID-19 epidemic was divided into four periods (Figure 1):

**Figure 1.**
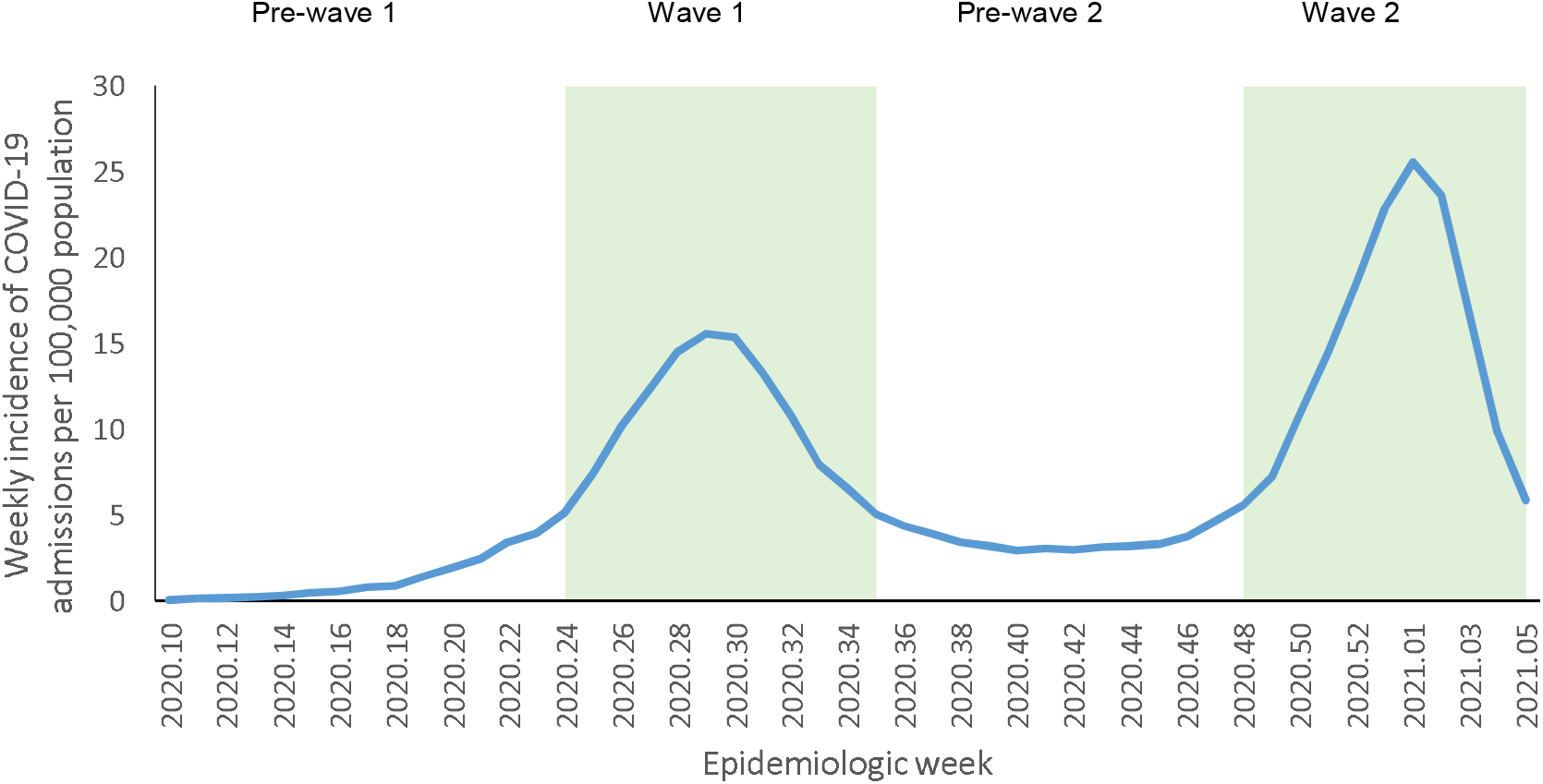
Weekly incidence per 100,000 population of COVID-19 admissions by epidemiologic week (shading shows the time periods of first and second waves), South Africa, 5 March 2020-6 February 2021, n=204,482.

- Pre-wave 1: weeks 10 – 23 (5 March – 6 June 2020)
- Wave 1: weeks 24 – 35 (7 June – 29 August 2020)
- Pre-wave 2: weeks 36 – 47 (30 August – 21 November 2020)
- Wave 2: week 48 of 2020 – week 5 of 2021 (22 November 2020 – 6 February 2021)

### Data analysis

We conducted two random effect multivariable logistic regression analyses. The first analysis assessed risk factors for in-hospital mortality accounting for wave period, and the second compared the characteristics of hospitalised COVID-19 cases in wave 1 and wave 2. For the multivariable model assessing risk factors for mortality, covariates included were age, sex, race, public or private health sector, presence of comorbidity, including wave period, and adjusting for weekly district COVID-19 admissions. In the mortality model, we included all four wave periods, *i*.*e*. pre-wave 1, wave 1, pre-wave 2, and wave 2. For the multivariable model comparing wave 1 and wave 2 as outcomes, covariates included were age, sex, race, health sector, presence of comorbidity and in-hospital mortality, also adjusting for weekly district COVID-19 admissions. The analysis included only data from wave 1 and wave 2, and not the other wave periods. Weekly COVID-19 numbers of admissions in the country were used as a proxy of burden of COVID-19 cases on the healthcare system and divided in four categories, low (<3,500), medium (3,500-7,999), high (8,000-12,499) and very high (>12,500). Only COVID-19 admissions were included as data on patients admitted with other illnesses were not available.

For all analyses we included all individuals with available data and included amongst the categories those who had missing data. A random effect on admission facility was included for all analyses to account for potential differences in the population served and the quality of care at each facility. For each multivariable model we assessed all variables that were significant at p<0.2 on univariate analysis and dropped non-significant factors (p≥0.05) with manual backward elimination. Pairwise interactions were assessed by inclusion of product terms for all variables remaining in the final multivariable additive model. Covariates included were chosen based on biological plausibility and evidence from previous analysis. Sensitivity analysis was done to separately assess the effect of including patients admitted in the private and public sectors.

We used the Chi squared test to assess the difference in CFR at the peak of the first and second wave. In addition, we compared the exponential growth rate of the first and the second wave. We estimated the exponential growth rate (from 5 admissions per 100,000 population at the start of the wave to the wave peak) for each wave using Poisson regression on count of weekly COVID-19 admissions (outcome variable) over time (weekly increases – dependent variable) and assessed the difference in the estimated weekly growth rate through the inclusion of an interaction term of weeks and wave in the model. The statistical analysis was implemented using Stata 15 (Stata Corp®, College Station, Texas, USA).

## RESULTS

### COVID-19 national trends

From 5 March 2020 to 6 February 2021, a total of 1,476,134 SARS-CoV-2 cases and 204,482 COVID-19 hospitalisations were reported in South Africa. Following the first wave peak in cases in epidemiologic week 28, there was a resurgence beginning in the Eastern Cape Province from week 40, followed by all other provinces subsequently, peaking in week 1 of 2021. Peak rates (per 100,000 persons) of COVID-19 cases, admissions and in-hospital deaths in the second wave exceeded the rates in the first wave (138.1 versus 240.1; 16.7 versus 28.9; and 3.3 versus 7.1 respectively (Figure 2).

**Figure 2.**
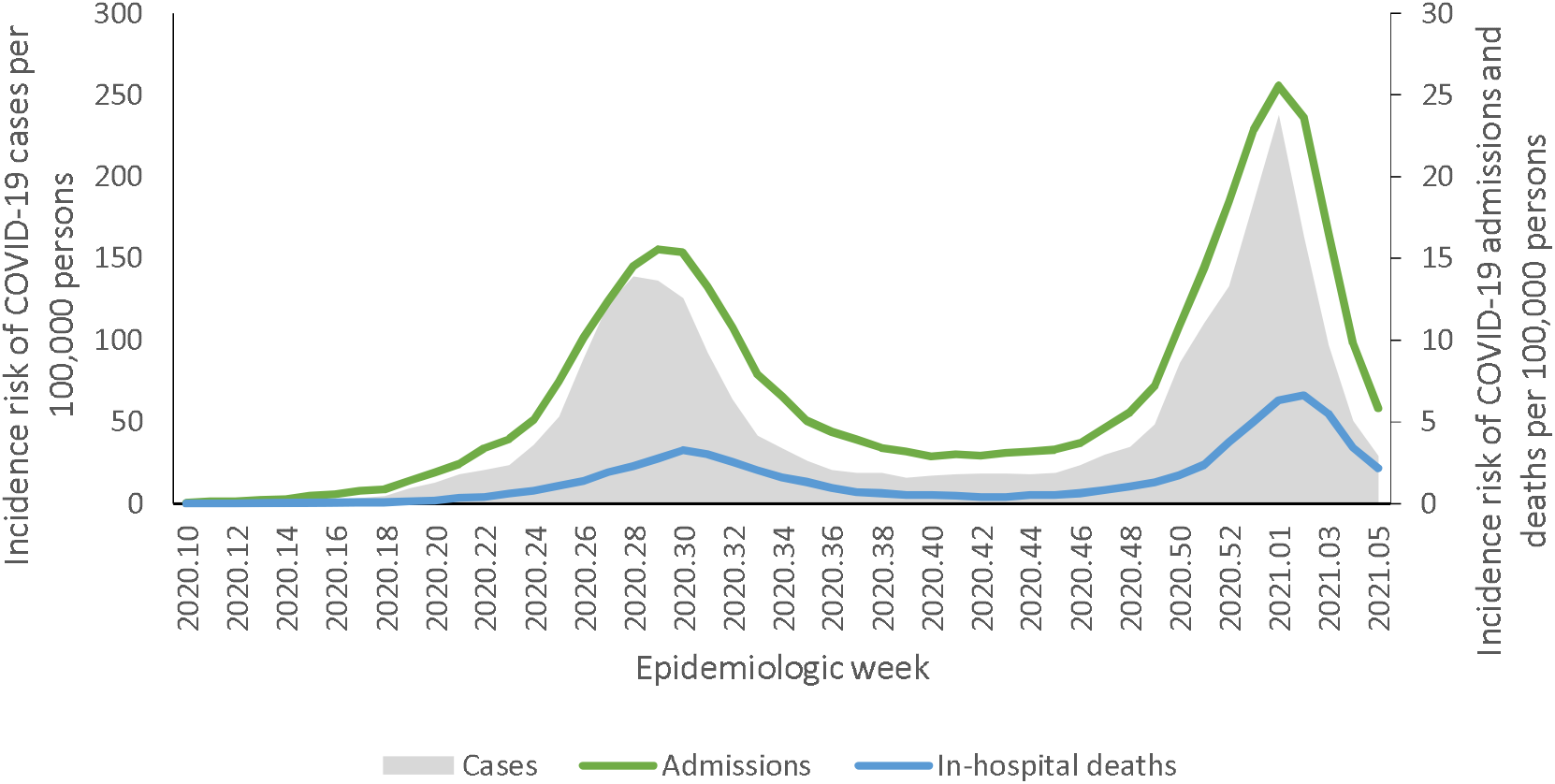
Incidence of reported SARS-Cov-2 cases, COVID-19 admissions and in-hospital deaths by epidemiologic week of diagnosis, South Africa, 5 March 2020-6 February 2021.

Of the 188,999 COVID-19 patients nationally with a recorded in-hospital outcome (died or discharged), 43,019 died and the in-hospital CFR was 22.8%. The CFR at the peak of the second wave in January (29.1%, 95% CI 28.6 – 29.5%) was significantly higher than the CFR at the peak of the first wave in July (20.8%, 95% CI 20.4 – 21.2%) (p<0.001) (Supplementary Table 1).

**Table 1.** Multivariable analysis of factors associated with in-hospital mortality among individuals hospitalised with COVID-19^*^, South Africa, 5 March 2020-6 February 2021.

| Characteristic | Case Fatality Ratio<br>n/N (%) | Unadjusted<br>OR | 95% CI | Adjusted OR | 95% CI | Adjusted OR | 95% CI |
| --- | --- | --- | --- | --- | --- | --- | --- |
|  |  |  |  | Including wave periods |  | Adjusting for weekly admissions |  |
| Age group (years) |  |  |  |  |  |  |  |
| <40 | 3040/45114 (6.7) | Reference |  | Reference |  | Reference |  |
| 40-64 | 20080/95181 (21.1) | **3.7** | 3.6-3.9 | **3.4** | 3.2-3.5 | **3.4** | 3.2-3.5 |
| ≥65 | 19754/47280 (41.8) | **9.9** | 9.5-10.3 | **8.8** | 8.4-9.2 | **8.7** | 8.3-9.1 |
| Unknown | 236/1460 (16.2) | **2.7** | 2.3-3.1 | **5.6** | 4.7-6.7 | **5.6** | 4.7-6.7 |
| Sex |  |  |  |  |  |  |  |
| Female | 22161/104962 (21.1) | Reference |  | Reference |  | Reference |  |
| Male | 20934/83908 (25.0) | **1.2** | 1.2-1.3 | **1.3** | 1.3-1.3 | **1.3** | 1.3-1.3 |
| Unknown | 15/164 (9.2) | **0.4** | 0.2-0.6 | **0.2** | 0.1-0.4 | **0.2** | 0.1-0.4 |
| Race |  |  |  |  |  |  |  |
| White | 2360/11112 (21.2) | Reference |  | Reference |  | Reference |  |
| Black | 22902/95000 (24.1) | **1.2** | 1.1-1.2 | **1.2** | 1.1-1.2 | **1.2** | 1.1-1.2 |
| Mixed | 2095/9119 (23.0) | **1.1** | 1.0-1.2 | **1.2** | 1.1-1.3 | **1.2** | 1.1-1.3 |
| Indian | 1673/7561 (22.1) | 1.1 | 0.9-1.1 | **1.2** | 1.1-1.3 | **1.2** | 1.1-1.3 |
| Other | 59/298 (19.8) | 0.9 | 0.7-1.2 | 1.0 | 0.7-1.3 | 1.0 | 0.7-1.3 |
| Unknown | 14021/65945 (21.3) | 1.0 | 0.9-1.1 | 1.1 | 0.9-1.1 | 1.1 | 0.9-1.1 |
| Comorbid condition |  |  |  |  |  |  |  |
| No co-morbidity | 11185/66387 (16.9) | Reference |  | Reference |  | Reference |  |
| Co-morbid condition | 22843/77155 (29.6) | **2.1** | 2.0-2.1 | **1.5** | 1.5-1.6 | **1.5** | 1.5-1.6 |
| Unknown | 9082/45493 (20.0) | **1.2** | 1.2-1.3 | 1.0 | 0.9-1.1 | 1.0 | 0.9-1.1 |
| Health sector |  |  |  |  |  |  |  |
| Private sector | 17763/95342 (18.6) | Reference |  | Reference |  | Reference |  |
| Public sector | 25347/93693 (27.1) | 1.6 | 1.6-1.7 | **1.6** | 1.4-1.8 | **1.6** | 1.4-1.8 |
| Province |  |  |  |  |  |  |  |
| Western Cape | 9063/41339 (21.9) | Reference |  | Reference |  | Reference |  |
| Eastern Cape | 9015/27643 (32.6) | **1.7** | 1.7-1.8 | **1.9** | 1.5-2.3 | **1.9** | 1.5-2.4 |
| Free State | 2259/10222 (22.1) | 1.0 | 0.9-1.1 | 1.2 | 0.9-1.6 | 1.2 | 0.9-1.5 |
| Gauteng | 9720 (50362 (19.3)) | **0.9** | 0.8-0.9 | **1.0** | 0.8-1.3 | **1.0** | 0.8-1.3 |
| KwaZulu-Natal | 7822/34530 (22.7) | **1.0** | 1.0-1.1 | **1.4** | 1.1-1.8 | **1.4** | 1.1-1.8 |
| Limpopo | 1752/5996 (29.2) | **1.5** | 1.4-1.6 | **1.7** | 1.3-2.3 | **1.7** | 1.3-2.3 |
| Mpumalanga | 1630/6516 (25.0) | **1.2** | 1.1-1.3 | **1.9** | 1.4-2.5 | **1.8** | 1.3-2.5 |
| North West | 1239/9408 (13.2) | **0.5** | 0.5-0.6 | 0.9 | 0.6-1.3 | 0.9 | 0.6-1.3 |
| Northern Cape | 610/3019 (20.2) | **0.9** | 0.8-0.9 | **1.6** | 1.0-2.4 | **1.6** | 1.0-2.3 |
| Wave period |  |  |  |  |  |  |  |
| Pre-wave 1 | 1710/9627 (17.8) | **0.8** | 0.8-0.9 | **0.9** | 0.8-0.9 | 1.0 | 0.9-1.0 |
| Wave 1 | 14669/72005 (20.4) | Reference |  | Reference |  | Reference |  |
| Pre-wave 2 | 3936/23428 (16.8) | **0.8** | 0.8-0.8 | **0.8** | 0.7-0.8 | **0.9** | 0.8-0.9 |
| Wave 2 | 22783/83913 (27.2) | **1.5** | 1.4-1.5 | **1.4** | 1.4-1.4 | **1.2** | 1.2-1.3 |
| Weekly national admission number |  |  |  |  |  |  |  |
| Low <3500 | 7706/43144 (17.9) | Reference |  |  |  | Reference |  |
| Medium 3500-7999 | 11264/53859 (20.9) | **1.2** | 1.2-1.3 |  |  | **1.1** | 1.0-1.2 |
| High 8000-12499 | 12629/53108 (23.8) | **1.4** | 1.4-1.5 |  |  | **1.2** | 1.2-1.3 |
| Very high >12500 | 11499/38862 (29.6) | **1.9** | 1.9-2.0 |  |  | **1.5** | 1.4-1.5 |

\* Statistically significant estimates are highlighted in bold OR=Odds Ratio; CI=Confidence Interval

### Rate of increase between the first and second wave

The time it took from 5 admissions per 100,000 to 15 admissions per 100,000 population in the first wave was six weeks and in the second wave was four weeks (Figure 3). The estimated weekly growth rate from the start to the peak of the first wave was 1.22 (95% CI 1.21-1.23; 22% weekly average incidence risk increase from week to week); and in wave 2 was 1.28 (95% CI 1.28-1.29; 28% weekly average incidence risk increase from week to week). There was a significantly higher rate of increase in wave 2 [ratio of growth rate in wave two compared to wave one:1.04, 95% CI 1.04-1.05].

**Figure 3:**
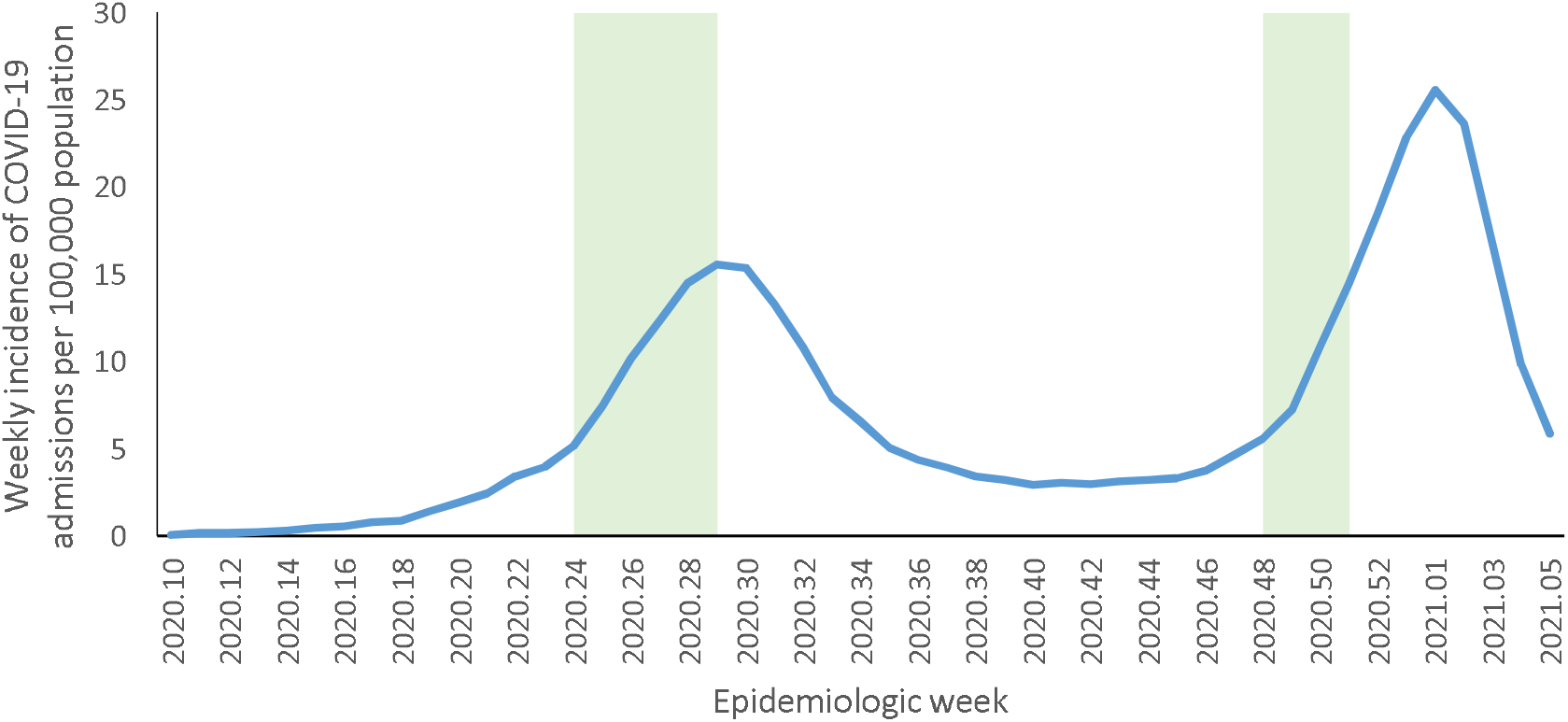
Incidence per 100,000 population of COVID-19 admissions by epidemiologic week (shading shows the time period from 5 admissions per 100,000 to 15 admissions per 100,000 in the first and second waves), South Africa, 5 March 2020-6 February 2021, n=204,482.

### In-hospital mortality in first and second wave

On multivariable analysis, among hospitalised individuals after adjusting for weekly hospital admissions, factors associated with in-hospital mortality were wave 2 [aOR 1.2, 95% CI (1.2 – 1.3)] and pre-wave 2 period [aOR 0.9, 95% CI (0.8 – 0.9)] compared to wave 1; age 40-64 years [aOR 3.4, 95% CI (3.2 – 3.5)], and >60 years [aOR 8.7, 95% CI (8.3 – 9.1)], compared to <40 years; male sex [aOR 1.3, 95% CI (1.3 – 1.3)]; Black [aOR 1.2, 95% CI (1.1 – 1.2)], mixed [aOR 1.2, 95% CI (1.1 – 1.3)] and Indian [aOR 1.2, 95% CI (1.1 – 1.3)] race compared to White race; presence of a comorbid condition [aOR 1.5, 95% CI (1.5 – 1.6)]; and admission in the public sector [aOR 1.6, 95% CI (1.4 – 1.8)]. Compared to weeks with low numbers of district hospital admissions, there was an increased risk of mortality in weeks with medium level of weekly admissions [aOR 1.1, 95% CI (1.0 – 1.2)], high weekly admissions [aOR 1.2, 95% CI (1.2 – 1.3)] and very high weekly admissions [aOR 1.5, 95% CI (1.4 – 1.5)]. We also observed differences in mortality by provinces (Table 1).

### Comparison of first and second wave

Multivariable analysis, comparing characteristics of hospitalised individuals in the first and second waves, after adjusting for weekly district hospital admissions, shows that there was an increase in in-hospital mortality in the second wave [aOR 1.2, 95% CI (1.2 – 1.3)]. The factors more common in the second wave were age 40-64 years [aOR 1.1, 95% CI (1.0 – 1.1)], and >60 years [aOR 1.1, 95% CI (1.1 – 1.1)], compared to <40 years; admission in the public sector [aOR 2.2, 95% CI (1.7 – 2.8)]; and high weekly admissions [aOR 1.6, 95% CI (1.6 – 1.7)] and very high weekly admissions [aOR 130661.3, 95% CI (8174 – 2088515)] compared to low weekly admissions. The factors less common in the second wave were admissions in individuals of Black [aOR 0.6, 95% CI (0.6 – 0.7)] and Indian [aOR 0.7, 95% CI (0.7 – 0.8)] race compared to White race; presence of a comorbid condition [aOR 0.5, 95% CI (0.5 – 0.5)]; and medium level of weekly admissions [aOR = 0.5, 95% CI (0.5 – 0.6)] compared to low weekly admissions. We also observed provincial differences between the first and second wave (Table 2).

**Table 2.**
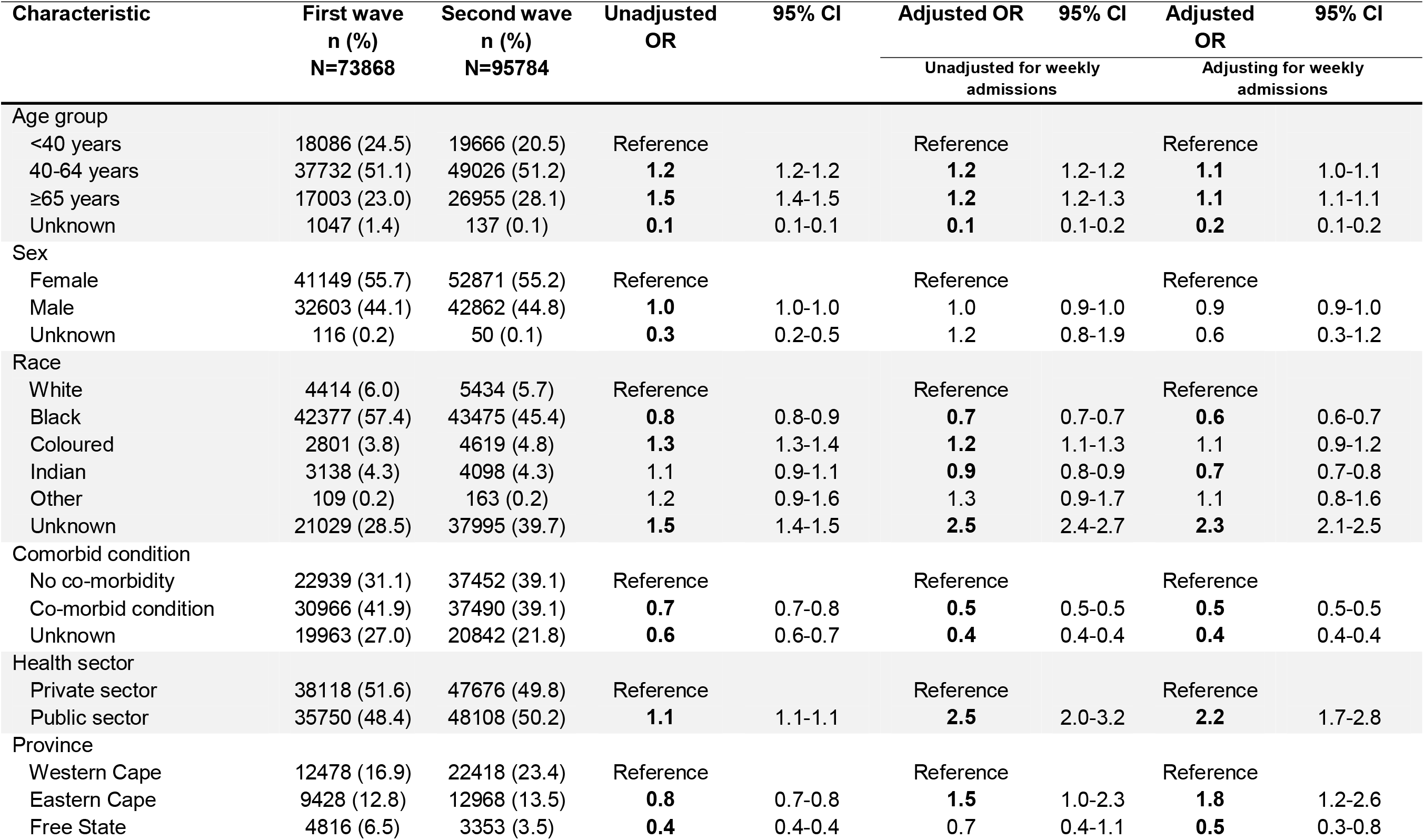
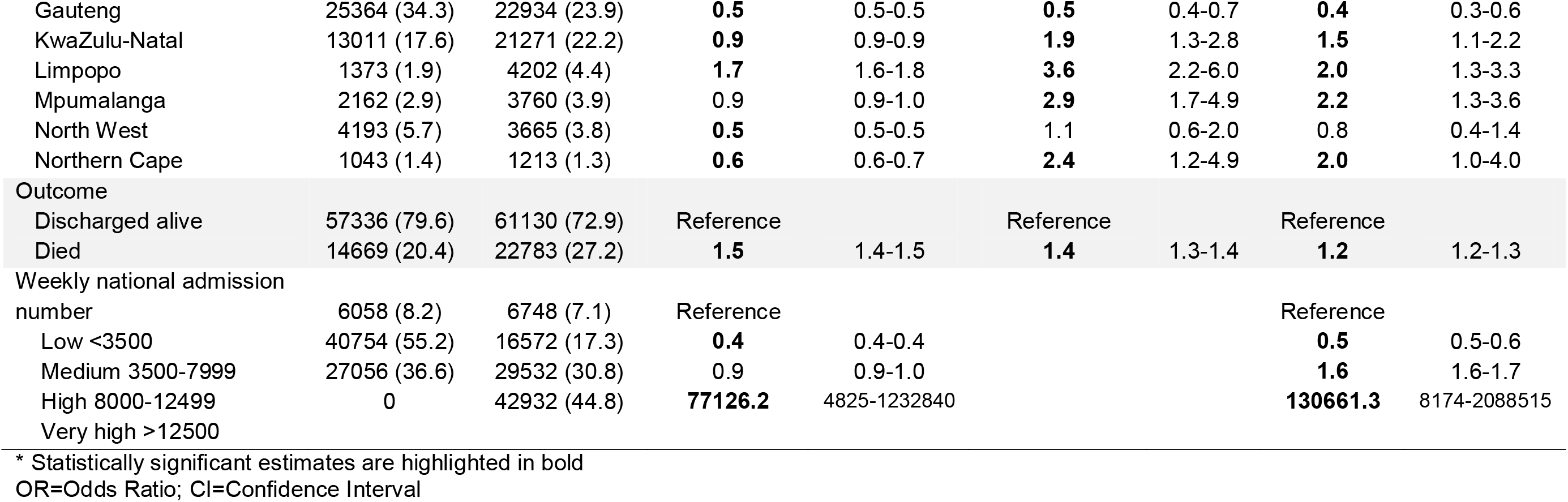
Multivariable analysis of factors associated with the second compared to the first COVID-19 wave among hospitalised individuals^*^, South Africa, 5 March 2020-6 February 2021.

In sensitivity analyses, we also separately analysed predictors of in-hospital death in the private sector (supplementary Table 2) and public sector (supplementary Table 3). Both comparisons showed increased mortality in the second wave compared to the first wave, and very similar trends and associations to those seen in the main combined analyses. Differences between sectors were higher estimates of association of second wave with mortality in the private sector (aOR 1.3, 95% CI 1.2-1.3) compared to the public sector (aOR 1.1, 95% CI 1.1-1.2); higher estimates of association of weekly admissions with mortality in the private sector; and association of race with mortality being present in the public sector and absent in the private sector.

## DISCUSSION

The incidence of COVID-19 cases, admissions and in-hospital deaths in the second wave exceeded the incidence in the first wave in South Africa. There was also a faster rate of increase of weekly incidence for COVID-19 admissions in the second wave and a 20% increase in in-hospital mortality in the second wave, even after adjusting for other factors. While we did not have individual-level data on infecting lineage for cases included in this analysis, the fact that 501Y.V2 has been the predominant lineage in the second wave (3,4) suggests that there may possibly be increased mortality associated with the new lineage. This is consistent with preliminary findings in the United Kingdom of increased CFR for individuals infected with variant B.1.1.7, with mortality hazard ratio estimates ranging from 1.35 to 1.91 in matched case-control and cohort analyses. (18)

Based on global trends, it might have been expected that in-hospital COVID-19 mortality in South Africa would have decreased in the second wave. In most countries, the second wave of COVID-19 had a higher number of cases but lower mortality (19-24). Improved outcomes during the second wave were likely a result of introduction of interventions such as remdesivir (25), dexamethasone (26), high-flow oxygen (27), greater use of thromboprophylaxis (28) as well as non-pharmacologic treatments such as placing the patient in the prone position.(20) Other possible suggestions for the lower CFR observed in the second wave in many countries are changes in demographic characteristics of cases, and the cohort or harvest effect with a large number of the elderly and those with health conditions (the vulnerable groups) likely to have died in the first wave (22,29,30). Additionally, improved testing capacities in the second wave could have resulted in more mild cases being identified (22,23,31,32); and healthcare systems in many countries could have been better prepared in the second wave, offering timely treatment of severe cases (20,22,30). Decreased mortality could also be related to the greater use of masks which, theoretically, could reduce the viral inoculum and perhaps disease severity (33).

The shifts in trend of admissions and deaths between wave 1 and 2 in South Africa could also be explained by the different context of the epidemic when compared to other countries. South Africa experienced the first wave two months later than other countries and benefitted from time for hospital preparedness and learning from other countries’ experiences and the use of steroids (34). The country was less well prepared for the second wave which was not predicted to have started as early as it did. Also South Africa instituted a hard lockdown for four weeks during wave 1, but very soft levels of restrictions during wave 2 due to concerns regarding the impact on people’s livelihoods and the economy.

On multivariable analysis, there was increased risk for mortality with admission load in South Africa. In weeks with very high weekly admissions, mortality increased by 50%, compared to weeks with low weekly district admissions. The observed increase in mortality of hospitalized patients at the peaks of the first and second waves reflects in part, increasing pressure on the health system. In South Africa, the rising number of hospitalisations in the second wave required care to be rationed to those patients highest on triage lists.(11) Studies have shown that a strain on hospital capacity has been associated with increased mortality in non-pandemic settings.(35) COVID-19 mortality in hospitals seems to be higher when the incidence of COVID-19 in the community was high or increasing (36,37) and when the number of hospitalisations were highest.(19) In addition, the rapid escalation in cases resulted in hospital resource constraints affecting outcomes.(38) Furthermore, strains on critical care capacity were associated with increased COVID-19 mortality.(38,39) In Brazil, a strained health-care system with regional differences in access to resources, compounded by overburdened hospital systems, contributed to greater in-hospital mortality.(40) It has been suggested that even the perception of a strained health system can lead to excess mortality from COVID-19 and other conditions, because individuals might avoid seeking care until their clinical condition has deteriorated or might die at home.(41) An important focus of the COVID-19 response in preparation for the third wave should be efforts to strengthen health system readiness and prepare hospitals and critical care services with additional surge capacity.

In the second wave, individuals who had comorbidities and were of Black or Indian race were less likely to be admitted to hospital in comparison to the first wave; while individuals who were older, and regular users of the public sector were more likely to be admitted. Across Europe, North America, the Middle East and South-East Asia, a shift towards younger cases with fewer comorbidities and less severe disease has been observed, considered to be due to public health measures to reduce transmission in vulnerable groups. (22,23,30,42,43,44) The higher proportion of older people admitted in the second wave in South Africa could be due to changes in preventive behaviour and transmission dynamics or increased susceptibility to the new lineage. The lower proportion of reported comorbidities in the second wave, even accounting for age distribution, could reflect differences in clinician practice, survival bias, or changing manifestation in individuals without underlying illness. It could also be due to variation in reporting of comorbidities, with under-ascertainment of other medical conditions at the peak of the wave being more likely when hospitals were busy.

Differences in race could reflect Black individuals being more greatly affected in the first wave due to historical differences in socio-economic status and housing conditions, which is supported by recent data showing higher sero-prevalence in Black individuals compared to other race groups in South Africa.(45) Some explanations for higher transmission and mortality in Black and Asian race groups have been suggested in the literature, including their over-representation in frontline occupations, higher incidences of multigenerational households, differences in access to health care, and public health messaging regarding prevention, early diagnosis, and treatment of COVID-19 being less effective in these groups, resulting in later presentation.(19) Our analysis showing that increased mortality is not seen by race group in the private sector, suggests that this finding may be related to health care access rather than any genetic or biological causes.

Provincial level differences may reflect differences in testing, health seeking behaviour, health systems, clinical practice and underlying population characteristics. Such regional differences have also been observed in other countries such as Brazil. (40)

It is important to note that even after adjusting for older age, higher admissions in the public sector, and higher hospital loads, there was still a significant independent increase in in-hospital mortality, which was not accounted for by other factors. However, our analysis differs from that of the United Kingdom who conducted a series of matched case control and population cohort studies comparing variant and non-variant cases. (18) We do not have individual level lineage type data, in part because there is no PCR-based test to distinguish the South Africa lineage as there is in the United Kingdom.

### Strengths and Limitations

Strengths of this study include the use of comprehensive electronic health record data on all COVID-19 hospitalizations at all 641 public and private hospitals in the country, minimizing selection or surveillance bias and maximising generalizability. The study includes a diverse patient population, complete study outcomes, and a lengthy period of investigation of 11 months, with in-hospital follow-up until occurrence of discharge or death.

Major limitations of this analysis were the lack of individual level lineage type data, and possible residual confounding as we could not adjust for several factors. We adjusted for COVID-19 admissions but were not able to adjust for weekly hospital admission volumes for persons under investigation (PUIs) and non-COVID-19 admissions. We are not able to adjust for differences between the first and second wave related to the level of national restrictions/lockdowns, and to individual preventive behaviours. The analysis includes only in-hospital deaths and any differences between the two waves in the proportions of patients who did not or could not access care and those that died outside of hospital are not accounted for in the analysis. There have been changes in treatment protocols with better COVID-19 treatment regimens including steroid use and high flow oxygen. These have likely decreased mortality rates as the epidemic progressed. The numbers of hospitals reporting to DATCOV increased in October 2020 and while all hospitals were required to back-capture historic admissions, they may not have done this completely, leading to reporting bias with possible underreporting in the first wave. The characteristics of those patients who died in the first wave (such as old age, comorbidities, obesity etc.) may have differed from the survivors and those who escaped infection. Therefore, the characteristics of those patients admitted to hospital in the second wave may be different compared to those of the first wave as a result of survival bias. Thresholds for hospital admission may also have changed over time.

## Conclusion

Our data suggest that the new lineage (501Y.V2) in South Africa may be one of the factors associated with increased in-hospital mortality during the second wave. Our data should be interpreted with caution however as our analysis is based on a comparison of mortality in the first and second wave as a proxy for dominant lineage and we did not have individual level data on lineage. Individual level studies comparing outcomes of people with and without the new lineage based on sequencing data are needed. To prevent high mortality in a potential third wave, we advocate a ‘flattening the curve’ approach, which requires a combination of strategies to slow the spread of COVID-19, to spread out the peak of the epidemic, which would prevent hospital capacity from being breached. (31)

## Data Availability

Data is currently shared in line with global COVID-19 hospital reporting, with the World Health Organization and the ISARIC, and will be made available on request.

## ACKNOWLEDGEMENTS

Thanks to all public and private sector hospitals and hospital groups submitting data to DATCOV, the National Department of Health for implementation support, the National Institute for Communicable Diseases for support and oversight, and the DATCOV team. We also acknowledge the Network for Genomics Surveillance in South Africa (NGS-SA) for sequence frequencies, and laboratories and clinicians throughout the country reporting cases and hospitalisation data.

## TABLE AND FIGURES

**Supplementary Table S1.** COVID-19 in-hospital case fatality ratio reported by month, South Africa, 5 March 2020-6 February 2021.

| Month | Died | Total outcomes | CFR (%) | 95% CI |
| --- | --- | --- | --- | --- |
| March | 9 | 247 | 3.6 | 1.7 – 6.8 |
| April | 113 | 963 | 11.7 | 9.8 – 13.9 |
| May | 710 | 3942 | 18.0 | 16.8 – 19.2 |
| June | 2680 | 13133 | 20.4 | 19.7 – 21.1 |
| July | 7228 | 34749 | 20.8 | 20.4 – 21.2 |
| August | 4891 | 24531 | 19.9 | 19.4 – 20.4 |
| September | 1696 | 10575 | 16.0 | 15.3 – 16.8 |
| October | 1142 | 8178 | 14.0 | 13.2 – 14.7 |
| November | 2018 | 9261 | 21.8 | 21.0 – 22.6 |
| December | 7351 | 29299 | 25.1 | 24.6 – 25.6 |
| January | 14125 | 48620 | 29.1 | 28.6 – 29.5 |

**Supplementary Table S2.** Multivariable analysis of factors associated with in-hospital mortality among individuals hospitalised with COVID-19, in private hospitals^*^, South Africa, 5 March 2020-6 February 2021.

| Characteristic | Case Fatality Ratio<br>n/N (%) | Unadjusted<br>OR | 95% CI | Adjusted OR | 95% CI | Adjusted OR | 95% CI |
| --- | --- | --- | --- | --- | --- | --- | --- |
|  |  |  |  | Including wave periods |  | Adjusting for weekly admissions |  |
| Age group |  |  |  |  |  |  |  |
| <40 years | 867/20204 (4.3) | Reference |  | Reference |  | Reference |  |
| 40-64 years | 8918/52657 (16.9) | **4.5** | 4.2-4.9 | **3.8** | 3.5-4.1 | **3.7** | 3.5-4.0 |
| ≥65 years | 7969/22433 (35.5) | **12.3** | 11.4-13.2 | **10.7** | 9.9-11.6 | **10.6** | 9.8-11.4 |
| Unknown | 9/48 (18.8) | **5.1** | 2.5-10.7 | **4.2** | 2.0-8.9 | **4.5** | 2.1-9.6 |
| Sex |  |  |  |  |  |  |  |
| Female | 8115/50157 (16.2) | Reference |  | Reference |  | Reference |  |
| Male | 9648/45183 (21.4) | **1.4** | 1.4-1.5 | **1.3** | 1.3-1.4 | **1.3** | 1.3-1.4 |
| Unknown | 0/2 (0) | - | - | - | - | - | - |
| Race |  |  |  |  |  |  |  |
| White | 1873/9479 (19.8) | Reference |  | Reference |  | Reference |  |
| Black | 5756/36124 (15.9) | **0.8** | 0.7-0.8 | **1.3** | 1.2-1.3 | **1.2** | 1.2-1.3 |
| Coloured | 922/4941 (18.7) | 0.9 | 0.9-1.0 | **1.3** | 1.2-1.5 | **1.3** | 1.2-1.4 |
| Indian | 1273/6267 (20.3) | 1.0 | 0.9-1.1 | **1.3** | 1.2-1.4 | **1.3** | 1.2-1.4 |
| Other | 9/45 (20.0) | 1.0 | 0.5-2.1 | 1.4 | 0.6-3.1 | 1.3 | 0.6-2.9 |
| Unknown | 7930/38486 (20.6) | 1.1 | 0.9-1.1 | **1.3** | 1.2-1.4 | **1.3** | 1.2-1.4 |
| Comorbid condition |  |  |  |  |  |  |  |
| No co-morbidity | 8622/53608 (16.1) | Reference |  | Reference |  | Reference |  |
| Co-morbid condition | 7986/33145 (24.1) | **1.7** | 1.6-1.7 | **1.6** | 1.5-1.7 | **1.6** | 1.5-1.7 |
| Unknown | 1155/8589 (13.5) | **0.8** | 0.8-0.9 | 0.9 | 0.8-1.0 | 0.9 | 0.8-1.0 |
| Province |  |  |  |  |  |  |  |
| Western Cape | 2877/16001 (18.0) | Reference |  | Reference |  | Reference |  |
| Eastern Cape | 2208/8589 (25.7) | **1.6** | 1.5-1.7 | **1.8** | 1.4-2.3 | **1.8** | 1.4-2.3 |
| Free State | 902/5101 (17.7) | 0.9 | 0.9-1.1 | **1.4** | 1.1-1.7 | **1.3** | 1.0-1.7 |
| Gauteng | 4987/29554 (16.9) | **0.9** | 0.9-0.9 | 1.1 | 0.9-1.3 | 1.1 | 0.9-1.2 |
| KwaZulu-Natal | 4522/22825 (19.8) | **1.1** | 1.1-1.2 | **1.3** | 1.1-1.6 | **1.3** | 1.1-1.5 |
| Limpopo | 849/3429 (24.8) | **1.5** | 1.4-1.6 | **1.6** | 1.2-2.2 | **1.6** | 1.2-2.1 |
| Mpumalanga | 560/3672 (15.3) | **0.8** | 0.7-0.9 | 1.1 | 0.9-1.5 | 1.1 | 0.8-1.5 |
| North West | 592/4403 (13.5) | **0.7** | 0.6-0.8 | 0.9 | 0.7-1.2 | 0.9 | 0.7-1.1 |
| Northern Cape | 266/1768 (15.1) | **0.8** | 0.7-0.9 | 1.2 | 0.8-1.7 | 1.1 | 0.8-1.7 |
| Wave period |  |  |  |  |  |  |  |
| Pre-wave 1 | 577/3796 (15.2) | 0.9 | 0.8-1.0 | 1.0 | 0.9-1.1 | **1.2** | 1.1-1.4 |
| Wave 1 | 6116/37507 (16.3) | Reference |  | Reference |  | Reference |  |
| Pre-wave 2 | 1405/11212 (12.5) | **0.7** | 0.7-0.8 | **0.7** | 0.7-0.8 | **0.9** | 0.8-0.9 |
| Wave 2 | 9658/42792 (22.6) | **1.5** | 1.4-1.6 | **1.5** | 1.5-1.6 | **1.3** | 1.2-1.3 |
| Weekly national admissions number | 2674/19720 (13.6) | Reference |  |  |  | Reference |  |
| Low <3500 | 4455/27142 (16.4) | **1.3** | 1.2-1.3 |  |  | **1.2** | 1.1-1.3 |
| Medium 3500-7999 | 5452/28181 (19.4) | **1.5** | 1.5-1.6 |  |  | **1.4** | 1.3-1.5 |
| High 8000-12499 | 5175/20264 (25.5) | **2.2** | 2.1-2.3 |  |  | **1.8** | 1.6-1.9 |
| Very high >12500 |  |  |  |  |  |  |  |

\* Statistically significant estimates are highlighted in bold OR=Odds Ratio; CI=Confidence Interval

**Supplementary Table S3.** Multivariable analysis of factors associated with in-hospital mortality among individuals hospitalised with COVID-19, in public hospitals^*^, South Africa, 5 March 2020-6 February 2021.

| Characteristic | Case Fatality Ratio<br>n/N (%) | Unadjusted<br>OR | 95% CI | Adjusted OR | 95% CI | Adjusted OR | 95% CI |
| --- | --- | --- | --- | --- | --- | --- | --- |
|  |  |  |  | Including wave periods |  | Adjusting for weekly admissions |  |
| Age group |  |  |  |  |  |  |  |
| <40 years | 2173/24910 (8.7) | Reference |  | Reference |  | Reference |  |
| 40-64 years | 11162/42524 (26.3) | **3.7** | 3.5-3.9 | **3.3** | 3.1-3.4 | **3.2** | 3.1-3.4 |
| ≥65 years | 11785/24847 (47.4) | **9.4** | 9.0-9.9 | **7.8** | 7.4-8.2 | **7.8** | 7.4-8.2 |
| Unknown | 227/1412 (16.1) | **2.0** | 1.7-2.3 | **6.0** | 5.0-7.3 | **6.0** | 5.0-7.2 |
| Sex |  |  |  |  |  |  |  |
| Female | 14046/54805 (25.6) | Reference |  | Reference |  | Reference |  |
| Male | 11286/38725 (29.1) | **1.2** | 1.2-1.2 | **1.3** | 1.3-1.3 | **1.3** | 1.3-1.3 |
| Unknown | 15/162 (9.3) | **0.3** | 0.2-0.5 | **0.2** | 0.1-0.4 | **0.2** | 0.1-0.4 |
| Race |  |  |  |  |  |  |  |
| White | 487/1633 (29.8) | Reference |  | Reference |  | Reference |  |
| Black | 17146/58876 (29.1) | 0.9 | 0.9-1.1 | 1.0 | 0.9-1.2 | 1.0 | 0.9-1.2 |
| Coloured | 1173/4178 (28.1) | 0.9 | 0.8-1.0 | 1.1 | 0.9-1.2 | 1.1 | 0.9-1.2 |
| Indian | 400/1294 (30.9) | 1.1 | 0.9-1.2 | 1.1 | 0.9-1.4 | 1.1 | 0.9-1.4 |
| Other | 50/253 (19.8) | **0.6** | 0.4-0.8 | 0.8 | 0.6-1.2 | 0.8 | 0.6-1.2 |
| Unknown | 6091/27459 (22.2) | **0.7** | 0.6-0.7 | **0.8** | 0.7-0.9 | **0.8** | 0.7-0.9 |
| Comorbid condition |  |  |  |  |  |  |  |
| No co-morbidity | 2563/12779 (20.1) | Reference |  | Reference |  | Reference |  |
| Co-morbid condition | 14857/44010 (33.8) | **2.0** | 1.9-2.1 | **1.5** | 1.4-1.6 | **1.5** | 1.4-1.6 |
| Unknown | 7927/36904 (21.5) | **1.1** | 1.0-1.1 | 1.0 | 1.0-1.1 | 1.0 | 0.9-1.1 |
| Province |  |  |  |  |  |  |  |
| Western Cape | 6186/25338 (24.4) | Reference |  | Reference |  | Reference |  |
| Eastern Cape | 6807/19054 (35.7) | **1.7** | 1.7-1.8 | **1.7** | 1.2-2.4 | **1.7** | 1.2-2.4 |
| Free State | 1357/5121 (26.5) | **1.1** | 1.0-1.2 | 0.9 | 0.6-1.5 | 0.9 | 0.6-1.5 |
| Gauteng | 4733/20808 (22.8) | **0.9** | 0.9-0.9 | 0.8 | 0.6-1.3 | 0.8 | 0.5-1.3 |
| KwaZulu-Natal | 3300/11705 (28.2) | **1.2** | 1.2-1.3 | 1.3 | 0.9-1.9 | 1.3 | 0.9-1.9 |
| Limpopo | 903/2567 (35.2) | **1.7** | 1.5-1.8 | **1.6** | 1.0-2.4 | **1.6** | 1.0-2.4 |
| Mpumalanga | 1070/2844 (37.6) | **1.9** | 1.7-2.0 | **2.0** | 1.3-3.1 | **1.9** | 1.2-3.1 |
| North West | 647/5005 (12.9) | **0.5** | 0.4-0.5 | 0.8 | 0.5-1.5 | 0.8 | 0.4-1.5 |
| Northern Cape | 344/1251 (27.5) | **1.2** | 1.0-1.3 | 1.7 | 0.9-3.1 | 1.7 | 0.9-3.1 |
| Wave period |  |  |  |  |  |  |  |
| Pre-wave 1 | 1133/5831 (19.4) | **0.7** | 0.7-0.8 | **0.7** | 0.7-0.8 | **0.8** | 0.7-0.9 |
| Wave 1 | 8553/34498 (24.8) | Reference |  | Reference |  | Reference |  |
| Pre-wave 2 | 2531/12216 (20.7) | **0.8** | 0.8-0.8 | **0.8** | 0.8-0.9 | **0.9** | 0.8-0.9 |
| Wave 2 | 13125/41121 (31.9) | **1.4** | 1.4-1.5 | **1.2** | 1.2-1.3 | **1.1** | 1.1-1.2 |
| Weekly national admissions number | 5032/23424 (21.5) | Reference |  |  |  | Reference |  |
| Low <3500 | 6809/26717 (25.5) | **1.3** | 1.2-1.3 |  |  | 1.1 | 0.9-1.1 |
| Medium 3500-7999 | 7177/24927 (28.8) | **1.5** | 1.4-1.5 |  |  | **1.1** | 1.1-1.2 |
| High 8000-12499 | 6324/18598 (34.0) | **1.9** | 1.8-2.0 |  |  | **1.2** | 1.1-1.3 |
| Very high >12500 |  |  |  |  |  |  |  |

\* Statistically significant estimates are highlighted in bold OR=Odds Ratio; CI=Confidence Interval

